# The influence of human genetic variation on Epstein-Barr virus sequence diversity

**DOI:** 10.1101/2020.12.02.20242370

**Authors:** Sina Rüeger, Christian Hammer, Alexis Loetscher, Paul J McLaren, Dylan Lawless, Olivier Naret, Daniel P. Depledge, Sofia Morfopoulou, Judith Breuer, Evgeny Zdobnov, Jacques Fellay, Swiss HIV Cohort Study

## Abstract

Epstein-Barr virus (EBV) is one of the most common viruses latently infecting humans. Little is known about the impact of human genetic variation on the large inter-individual differences observed in response to EBV infection. To search for a potential imprint of host genomic variation on the EBV sequence, we jointly analyzed paired viral and human genomic data from 268 HIV-coinfected individuals with CD4+ T cell count <200/mm^3^ and elevated EBV viremia. We hypothesized that the reactivated virus circulating in these patients could carry sequence variants acquired during primary EBV infection, thereby providing a snapshot of early adaptation to the pressure exerted on EBV by the individual immune response. We searched for associations between host and pathogen genetic variants, taking into account human and EBV population structure. Our analyses revealed significant associations between human and EBV sequence variation. Three polymorphic regions in the human genome were found to be associated with EBV variation: one at the amino acid level (BRLF1:p.Lys316Glu); and two at the gene level (burden testing of rare variants in BALF5 and BBRF1). Our findings confirm that jointly analyzing host and pathogen genomes can identify sites of genomic interactions, which could help dissect pathogenic mechanisms and suggest new therapeutic avenues.

## Introduction

Human genetic variation plays a key role in determining individual responses after exposure to infectious agents. Even though susceptibility or resistance to a microbial challenge is the final result of dynamic interactions between host, pathogen, and environment, human genetic polymorphisms have been shown to have an important, directly quantifiable impact on the outcome of various infections [1,2].

Genome-wide association studies (GWAS) have proven powerful to identify genetic regions implicated in a wide range of complex traits in both health and disease [3]. In the field of infectious diseases, several clinical and laboratory phenotypes have been investigated, including, for example disease susceptibility [4,5], clinical outcomes [6], adaptive immunity [7,8,9] or drug response [10]. In chronically infected patients, however, the pathogen genome itself provides a promising complementary target to investigate the impact of host genomic diversity on infection. While one part of the variation observed in pathogen DNA or RNA sequence is present at the transmission event, another fraction is acquired during the course of an infection, resulting at least partially from selective pressure exerted by the host response on the infectious agent. The phenomenon of within-host evolution has been extensively investigated for both viruses [11,12,13] and bacteria [14,11]. Pathogen genomic variation can thus be considered an intermediate phenotype that is detectable as a footprint of within-host evolution. This can serve as a basis for a joint association analyses of host and pathogen genome variation, which we called genome-to-genome (G2G) analysis [15], a more powerful approach than using a clinical outcome alone. A global description of the adaptive forces acting on a pathogen genome during natural infection holds the potential to identify novel therapeutic and diagnostic targets and could inform vaccine design efforts [16].

A G2G analysis for the quickly evolving human immunodeficiency virus (HIV) identified strong associations of single nucleotide polymorphisms (SNPs) in the HLA class I region with multiple amino acid variants across the viral genome [15]. More recent work showed an impact of variation in the HLA class II and interferon lambda 4 (IFNL4) loci on hepatitis C virus (HCV) sequence diversity [17,18,19]. While the rate of evolutionary change in RNA viruses is higher than in DNA viruses [20], the latter also present considerable amounts of inter- and intra-host variation. Among herpesviruses, it has been shown that human cytomegalovirus (HCMV) has higher genomic variability than other DNA viruses [21]. Recent genome sequencing efforts demonstrated that the same holds true for Epstein-Barr virus (EBV) [22,23].

EBV is a widespread human pathogen that causes infectious mononucleosis in about 10% of individuals during primary infection. EBV infection occurs most often early in life, with about 30% of children being seropositive by age 5, 50% by age 10 and up to 80% by age 18 [24]. This human infecting herpesvirus has also been associated with post-transplant lymphoproliferative disease [25] and could play a role in some autoimmune diseases [26,27]. In addition, EBV has oncogenic properties and is implicated in the pathogenesis of multiple cancer types, predominantly Burkitt’s lymphoma, Hodgkin’s and non-Hodgkin’s lymphoma, nasopharyngeal carcinoma and gastric carcinoma [28,29]. More than 5% of the 2 million infection-associated new cancer cases in 2008 could be attributed to EBV [30]; it was also estimated to have caused 1.8% of cancer deaths in 2010, i.e. more than 140,000 cases [31].

The EBV genome is approximately 170 Kbp long and encodes at least 80 proteins, not all of which have been definitively identified or characterized. After primary infection, the EBV genome persists in B cells as multicopy episomes that replicate once per cell cycle. In this latent mode, only a small subset of viral genes is expressed. Latent EBV can then reactivate to a lytic cycle, which involves higher gene expression and genome amplification for packaging into new infectious viral particles [32].

A small number of host genomic analyses of EBV infection have been recently published, demonstrating that human genetic diversity plays a role in disease outcome. A study in 270 EBV isolates from southern China identified two non-synonymous EBV variants within the *BALF2* gene that were strongly associated with the risk of nasopharyngeal carcinoma [33]. Another group investigated the co-evolution of worldwide EBV strains [34] and found extensive linkage disequilibrium (LD) throughout EBV genomes. Furthermore, they observed that genes in strong LD were enriched in immunogenic genes, suggesting adaptive immune selection and epistasis. In a pediatric study of 58 Endemic Burkitt lymphoma cases and 40 healthy controls, an EBV genome GWAS identified 6 associated variants in the genes *EBNA1, EBNA2, BcLF1*, and *BARF1* [35]. Finally, the narrow-sense heritability of the humoral immune response against EBV was estimated to be 0.28 [9,36].

Here, we present the first global analysis of paired human and EBV genomes. We studied full EBV genomes together with their respective host genomic variation in a cohort of 268 immunocompromised, HIV-coinfected patients. We chose untreated HIV-coinfected patients because EBV reactivation leading to viremia is more prevalent in immunosuppressed individuals than in an average population. Our analysis reveals three novel host genomic loci that are associated with variation in EBV amino acids or genes.

## Materials and Methods

### Study participants, sample preparation

The Swiss HIV Cohort Study (SHCS) is a nationwide, prospective cohort study of HIV-infected patients that enrolled >20,000 individuals since its establishment in 1988 and prospectively followed them at 6-month intervals [37]. For this project, SHCS participants were identified based on written consent for human genetic testing and availability of a peripheral blood mononuclear cell (PBMC) sample at time of advanced immunosuppression (i.e., with CD4+ T cell count below 200/mm^3^) in the absence of antiretroviral treatment.

We obtained demographic and clinical information from the SHCS database. These included sex, age, longitudinal HIV viral load results (number of RNA copies per ml of plasma), longitudinal CD4+ T cell counts (number of cells per mm^3^ of blood), and history of opportunistic infections.

The SHCS has been approved by the Ethics Committees of all participating institutions (Ethikkommission Nordwest-und Zentralschweiz, EKNZ; Kantonale Ethikkommission Bern; Ethikkommission Ostschweiz, EKOS; Ethikkommission Zürich; Commission cantonale d’éthique de la recherche sur l’être humain, Genève, CCER; Commission cantonale d’éthique de la recherche sur l’être humain, Vaud, CER-VD; Comitato etico cantonale Ticino). Each study participant provided written informed consent for genetic testing, and all research was performed in accordance with relevant guidelines and regulations.

### EBV genome quantification, enrichment and sequencing

DNA was extracted from PBMCs using the MagNA Pure 96 DNA and the Viral NA Small Volume Kit (Roche, Basel, Switzerland). Cellular EBV load was then determined using quantitative real-time PCR. Samples that yielded > 100 viral copies / ul were selected for EBV genome sequencing.

We used the previously described enrichment procedure to increase the relative abundance of EBV compared to host DNA [38]. Shortly, baits covering the EBV type 1 and 2 reference genomes were used to selectively capture viral DNA according to the SureSelect Illumina paired-end sequencing library protocol. Samples were then multiplexed and sequenced on an Illumina NextSeq sequencer [38].

### EBV sequence analyses

We chose a reference-based approach to call variants in the pathogen data. Since EBNA-2 and EBNA-3s are highly variable between EBV-1 and EBV-2 strains, we suspected that reads sequenced from these genes would map only to their corresponding type. In an attempt to attenuate the reference-bias this could cause, we constructed two references, one with the whole genome of the EBV-1 strain B95-8 (accession NC_007605) and EBNA-2 and EBNA-3s sequences from EBV-2 strain AG876 (accession NC_009334) and another one with the whole genome of AG876 with the EBNA-2 and EBNA-3s sequences of B95-8.

The read libraries were processed through Trimmomatic [39] to remove remnant PCR tags, TagDust [40] to eliminate low complexity reads and CD-HIT [41] to filter out duplicate reads. The remaining sequence reads were aligned to the constructs described in the previous paragraph. Following GATK best practices [42,43], we mapped the read libraries using BWA mem [44]. We cleaned the regions around InDels using GATK v3.8’s IndelRealigner [45]. As a last pre-processing step, we applied bwa-postalt.js, a BWA script that adjusts mapping quality score in function of alignments on ALT haplotypes.

Because patients can be infected by multiple EBV strains [46], we used BWA’s ALT-aware ability. In short, reads mapping to an ALT contig were always marked as supplementary alignment, regardless of mapping quality, unless they did not map to the primary assembly. This makes it easy to find unambiguously mapped reads, which we used as markers to quantify type 1 and type 2 EBV reads in all samples.

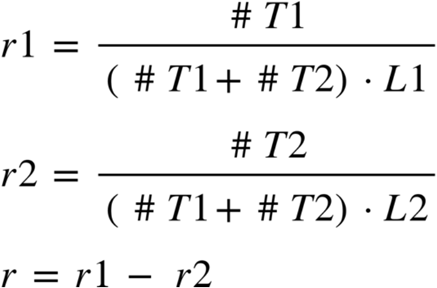

where *#T1* and *#T2* are the unambiguous read counts against type 1 and 2 haplotypes, respectively, *L1* and *L2* are the length of type 1 and 2 haplotypes, respectively, and *r1* and *r2* are the type 1 and 2 ratios, respectively. The score *r* is the relative abundance between type 1 and type 2.

### Definition of EBV amino acid variants

Since no gold standard variant set exists for EBV nor any closely related viral species, variant calling was performed using three different variant callers (GATK haplotypecaller, SNVer [47] and VarScan2 [48]) and by selecting as *bona fide* variant set the intersection of the three. The identified EBV variants were annotated using snpEff [49]. Nucleotide variants were transformed into binary amino acid matrices using in-house python scripts. The whole pipeline is written in Snakemake [50] and Python [51].

This approach was benchmarked using synthetic libraries generated from B95-8 and AG876 using ART Illumina and RNFtools, at a range of coverage between 10X and 250X and 5 different admixture conditions, 100% B95-8 or AG876, 75% - 25 % and 50%-50%. Assessing the true number of variants between EBV-1 and EBV-2 strains is not trivial because of the high variability in EBNA-2 and EBNA-3s regions. Therefore, we rated the variant callers and the consensus of the three mentioned callers on self-consistency. The performances of the runs were measured using the ratio of the variant counts to the size of the union of all variants called by a specific tested tool.

By using EBV type 2 as a reference, we focused on two types of variation in EBV strains: 1) single amino acid variants; and 2) burden of very rare amino acid variants (present in only 1 sample) in each viral gene (Figure 1). We call these datasets *EBV amino acids* and *EBV genes*, respectively. Both datasets contain binary values, with a value of 1 standing for “variant present” and 0 for “no variant present”. Positions with a coverage of less than 6x were set to “missing” and samples with more than 80% missing positions were excluded entirely. The positions covered by less than 6 reads were considered missing and imputed using the imputePCA function implemented in the missMDA R-package [52]. In total, we obtained 4392 amino acid variants and 83 gene variants. However, to limit the risk of model overfitting and because of low statistical power due to sample size we only included in the downstream association analyses the 575 *EBV amino acids* with an amino acid frequency of more than 10% and 52 *EBV genes*.

**Figure 1:**
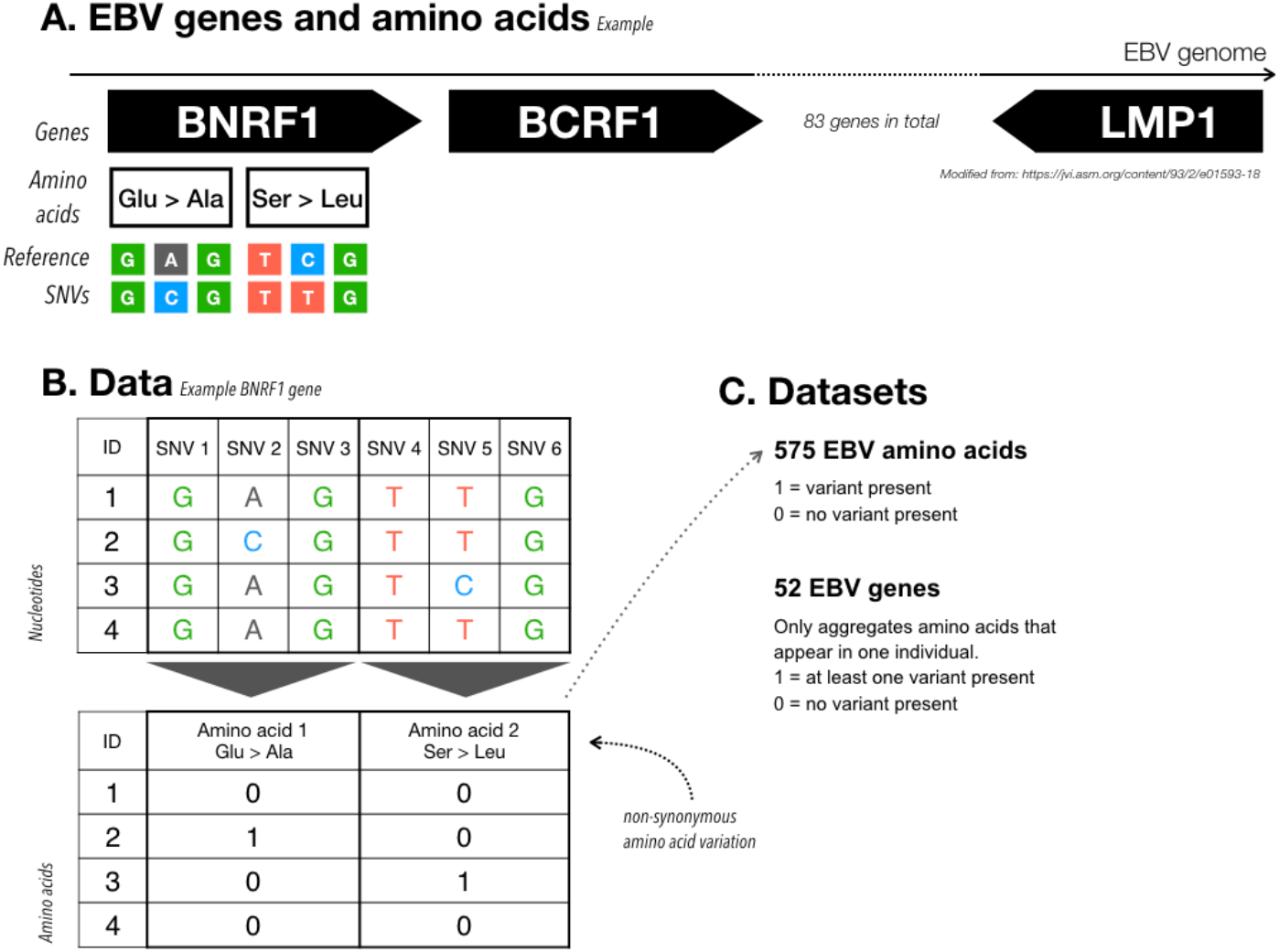
Illustration of EBV sequence variation. A) The EBV genome is about 170 Kbp long and contains 83 genes, for a total of 4392 amino acid residues. As an example, we focus on the *BNRF1* gene and on two amino acid changes: Glu→Ala and Ser→Leu. We know for each sample the genomic variants across the whole genome, as illustrated with the colored nucleotides. Using the nucleotide information and a reference genome we can compute the amino acid changes. B) We compare each individual (ID) to reference data and encode an amino acid as 1 if that individual has a non-synonymous change, and a 0 if not. This process returns us a matrix containing binary values, with individuals as row, and amino acids as columns. In our example, individual 2 has an amino acid change Glu→Ala and individual 3 an amino acid change Ser→Leu. C) To transform the data into outcomes for the G2G analysis we can use the amino acid matrix as it is (*EBV amino acids* dataset) or remove all amino acid columns that appear in more than 1 individual and then pool amino acids per gene (1 = variant present, *EBV genes* dataset).

### Human genotyping and imputation

A subset of 84 participants had been genotyped in the context of previous studies on several platforms. For the remaining 196 samples, human genomic DNA was isolated from PBMCs with the QIAsymphony DSP DNA Kit (Qiagen, Hilden, Germany), and genotyped using Illumina OmniExpress (v1.1) BeadChip arrays.

Genotype imputation was performed on the Sanger imputation server independently for all genotyping platforms, using EAGLE2 [53] for pre-phasing and PBWT [54] with the 1000 Genomes Phase 3 reference panel [55]. Low-quality imputed variants were excluded based on imputation INFO score (< 0.8). All datasets were merged, only keeping markers that were genotyped or imputed for all genotyping platforms. SNPs were excluded on the basis of per-individual missingness (> 3%), genotype missingness (> 1%), marked deviation from Hardy-Weinberg equilibrium (p < 1×10^−6^) and minor allele frequency < 5% (Table 1). All quality control procedures were performed using PLINK 2.0 [56].

**Table 1:** Summary of pathogen variants, host SNPs and covariates for 268 individuals. For each covariate, we indicate the number of individuals measured, and distribution (mean, median, standard deviation, minimum, maximum for quantitative, frequency for sex). For aggregated EBV genes the frequency is shown, for host SNPs the MAF distribution is presented.

| dataset | variable | counts | mean | median | sd | min | max |
| --- | --- | --- | --- | --- | --- | --- | --- |
| Pathogen genome (83 Rare EBV gene variation) | Variant frequency |  | 0.067 | 0.049 | 0.057 | 0 | 0.37 |
| Pathogen genome (575 EBV amino acids) | Variant frequency |  | 0.26 | 0.24 | 0.12 | 0.093 | 0.5 |
| Covariates | Sex | male: 206,<br>female: 62 |  |  |  |  |  |
| Covariates | AGE |  | 42.02 | 40.79 | 10.98 | 20.25 | 77.52 |
| Covariates | PC1 |  | -8.95 | -7.12 | 32.53 | -69.95 | 76.11 |
| Covariates | PC2 |  | 7.39 | 8.4 | 30.41 | -63.83 | 61.8 |
| Covariates | PC3 |  | 4.45 | 3.19 | 21.41 | -46.44 | 51.11 |
| Covariates | PC4 |  | -3.91 | -6.69 | 20.72 | -36.49 | 52.11 |
| Covariates | PC5 |  | -5.09 | -8.39 | 18.76 | -51.7 | 56.02 |
| Covariates | PC6 |  | -3.16 | -3.21 | 18.65 | -55.69 | 35.68 |
| Covariates | EBV type |  | 0.54 | 0.95 | 0.67 | -1 | 1 |

### Association analyses

We used the mixed model association implementation for binary and continuous outcomes in GCTA (v1.92) [57,58] to search for potential associations between human SNPs and EBV variants. The model can be expressed with the following equation:

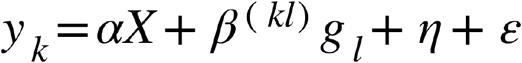

where the outcome y is a binary vector indicating whether an EBV variant is present (1) or not (0); X is a matrix that contains all covariates, α represents all fixed effects of all covariates (including an intercept term), g is the SNP genotype vector with coded additive allele dosages 0, 1 or 2, β is the (fixed) effect of the SNP to be tested for association, η is the polygenic (random) effect and ε the error term. This mixed model was estimated for each EBV variant *(k)* and SNP (*l)*, and integrated over all L SNPs and K EBV variants. To estimate η, the host genetic relationship matrix (GRM) was calculated from QC preprocessed genotype data using GCTA [57].

The use of a mixed effects association model allows to account for population stratification of the host genome. To control for population stratification among EBV genomes, we included the first six principal components (PCs) of EBV genetic variation to the covariate matrix X [59]. Other covariates were sex, age and EBV type. PCs were calculated from EBV amino acid variants using the convexLogisticPCA function from the R package logisticPCA [60] in R [61]. As data preparation for PC computation, we removed variants with less than 5% or more than 95% frequency. Missing amino acid values were imputed with the imputePCA function from the R package missMDA [52].

Significance was assessed using the usual genome-wide significance threshold in European populations of 5×10^−8^ and dividing it by the effective number of GWASs performed [62]. We used FINEMAP [63] to determine the most likely causal SNP(s) in a 2-Mb-wide window around each significant SNP. FINEMAP requires GWAS summary statistics and LD estimations as input. To estimate LD between SNPs, we used LDstore [64]. We performed eQTL lookups for host SNPs in eQTLGen [65], EUGENE [66] and GTEx [67].

Unless otherwise specified, all data preparation and analyses were performed using R [61].

### Code availability

EBV data preparation: https://gitlab.com/ezlab/vir_var_calling

G2G Analysis: https://github.com/sinarueeger/G2G-EBV-manuscript

## Results

### Study participants and human genetic data

PBMC samples from 778 SHCS participants were screened for the presence of cellular EBV DNA using RT-PCR. A total of 290 of them were identified as viremic for EBV (>2000 copies). We obtained good quality human genotyping and EBV sequencing data for 268 of them, which were included in the association analyses. The study cohort comprised 206 male and 62 female individuals, between the ages of 20 and 78 (median 40) (Table 1 and Supplementary Figure S1).

We applied standard GWAS quality control (QC) procedures that yielded information for 4’291’179 SNPs (Table 1 and Supplementary Figure S2, which shows the distribution of the minor allele frequency spectrum after QC).

### EBV genomic diversity and variant calling

Genome coverage was very uneven between the samples. Mean depth varied from less than 6x for 14 samples, up to more than 500x in 5 others. We also observed fluctuation in coverage above 6x, which we used to exclude 12 samples in which less than 20% of the EBV genome was sufficiently covered (Supplementary Figure S7a). In addition, the coverage in the first sequencing batch was not uniform.

We estimated the clonality of EBV strain in each sample by taking advantage of the high divergence between EBNAs T1 and T2 haplotypes. Among the 282 sequenced samples, 57.1% were predominantly (9:1) infected by T1 EBV, while 5.7% were mostly infected by T2 EBV (Supplementary Figure S6). The remaining 37.2% were infected by multiple strains or by recombinant viruses. This approach does not allow to stratify further than the EBNA types.

The variant calling pipeline was adapted to output variants by minimizing the impact of the admixture ratio and of the low coverage observed in the SHCS samples. Variants were called against EBV-2, as EBV-2 was able to call more variants than EBV-1 (Supplementary Figure S7d). The benchmark experiments against AG876 (EBV-2) yielded a total of 961 different variants. The most conservative was SNVer (783 variants), while the most sensitive was BCFtools (930 variants). The variant callers can be prone to artifacts [68], which was specifically observed in SNVer (Supplementary Figure S7c) in these datasets. To reduce the probability of calling artifacts, we chose to use the *bona fide* intersection of GATK HC, SNVer and VarScan2. This approach is likely to be impacted by low coverage. The recall is stable at around 95% at 25X coverage upwards and reasonable (10%) at 20X (Supplementary Figure S7c). Hence, low coverage has an impact, specifically, half potential variants called, on only 15% of the SHCS sample. However, this approach is very conservative, since it outputs only 88%, 85% and 79% of the variants called by SNVer, GATK HC and VarScan2, respectively.

On average, around 800 amino acid variants were called for each sample, with slight differences correlating with the clonality of the samples and the coverage above 6X (Supplementary Figure S7d). The variant counts against the AG876 construct (EBV-2) were generally higher in mixed infections and EBV-1 strains (Supplementary Figure S7d A). The variant counts were generally lower in the samples included in the first sequencing batch, which is likely due to the fluctuating coverage. However, overall, the number of variants was found to be comparable across the samples, ranging from 400 to 1500 for the 77% samples with a 6X coverage above 80% (Supplementary Figure S7d). Under 80% coverage, the variant counts hardly exceed 500 but rarely drops under 200 either. It is therefore likely that we missed variants using our approach. The positions covered by less than 6 reads were considered missing and imputed afterwards using the imputePCA function implemented in the missMDA R-package.

We analyzed EBV variation using two approaches: single marker analysis of *EBV amino acids, to investigate common viral variation*, and burden testing of very rare amino acid variants in *EBV genes* (Table 1, Supplementary Figure S3). Applying logistic principal component analysis of viral genomic structure showed a single main cluster (Supplementary Figure S4).

### Genome-to-Genome association analysis

We tested for associations between each EBV variant and human SNPs. We studied 575 EBV amino acids and 52 EBV genes, for a total of 627 GWASs. The effective number of GWASs performed was 458. As covariates, we included the first six EBV principal components (51.4% deviance explained), sex, age, type 1 vs 2 of EBV (Supplementary Figure S1). The sample size ranged between 120 and 268, with a median sample size of 264. Sample size variation was due to variable missingness in the EBV data. Genomic inflation factors for each of the 627 GWASs ranged between 0.92 and 1.12.

Significant associations (p < 1.09×10^−10^) were identified between a total of 25 human SNPs and viral variants mapping to three EBV regions (Table 2): the *EBV genes* **BALF5** (Figure 2a) and **BBRF1** (Figure 2b) and the *EBV amino acid* **BRLF1:p**.**Lys316Glu** (Figure 2c). The minor allele frequency of all significant host SNPs was between 0.05 and 0.10. The genomic inflation factors of the three GWASs ranged between 0.95 and 0.96 (Q-Q plots shown in Supplementary Figure S5).

**Figure 2:**
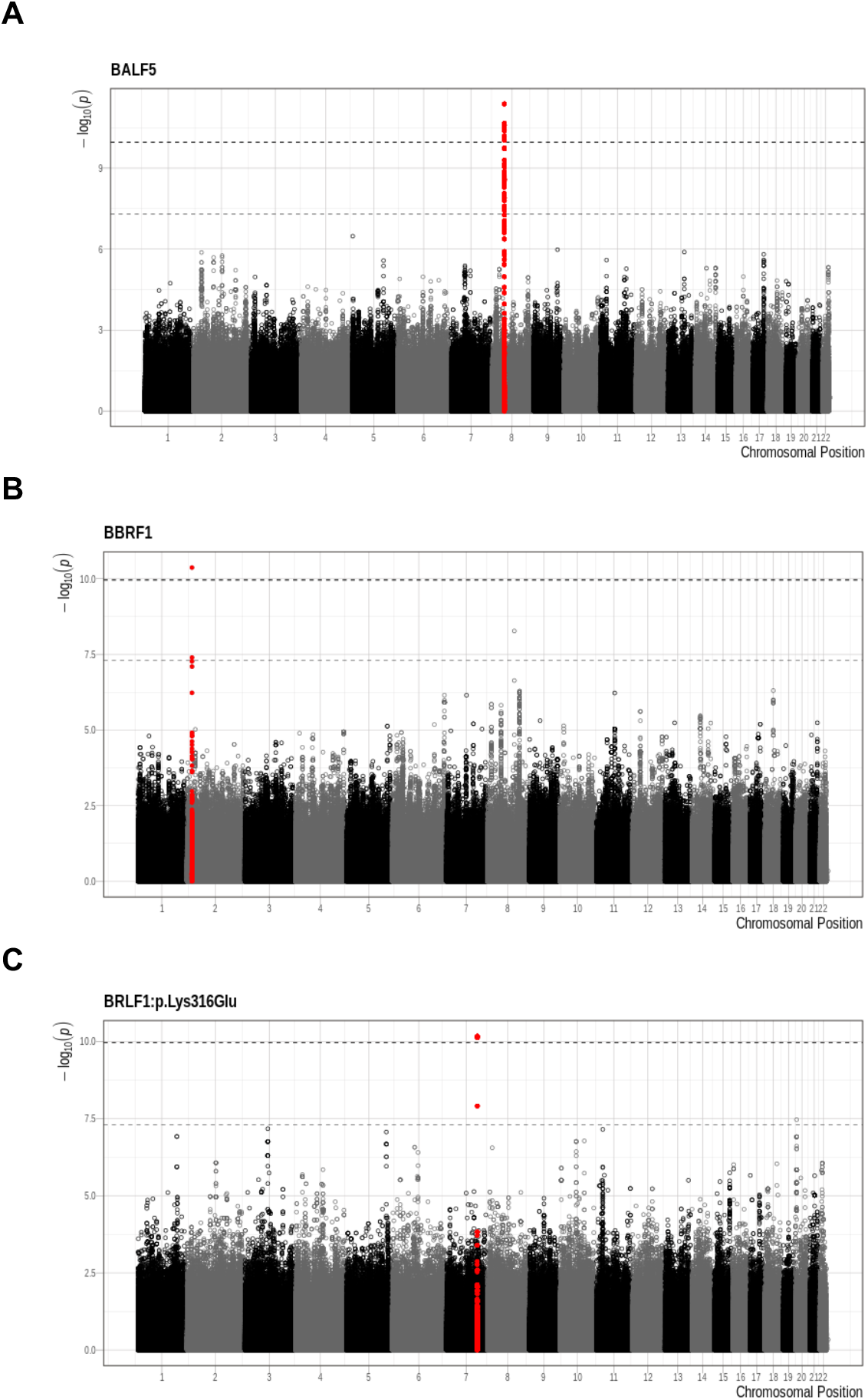
Significant associations - A: *BALF5*, B: *BBRF1*, C: *BRLF1*:p.Lys316Glu. The x-axis represents the chromosomal position and the y-axis displays the -log10(p-value). Colour alternates between chromosomes. Regions that contain statistically significant SNP are presented in red (top SNP +/- 400 Kbp). The light grey dashed line represents the GWAS significance threshold of 5×10^−8^, the dark grey dashed line the G2G threshold of 1.09×10^−10^.

**Table 2:**
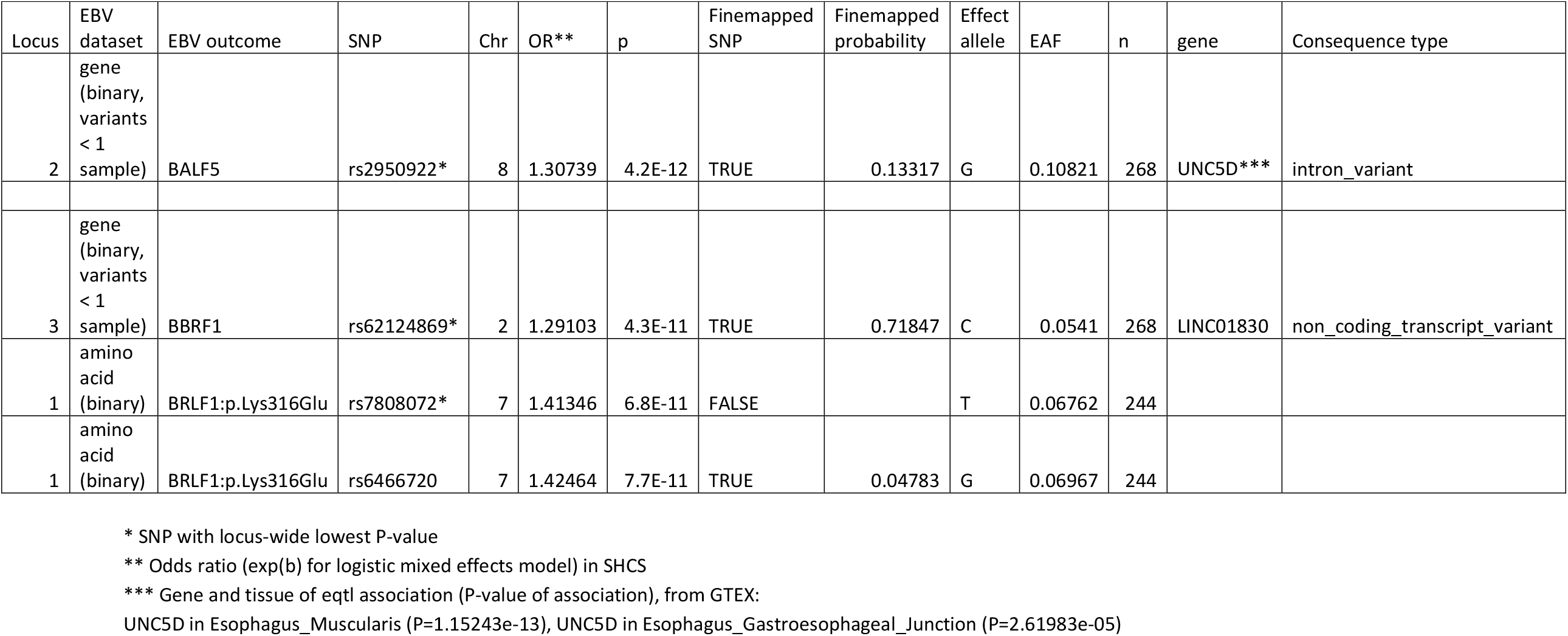
Summary of G2G analysis results. Top SNP and/or fine-mapped SNP per locus represented with: EBV dataset, EBV outcome, chromosome, SNP identifier, odds ratio, p-value, whether this SNP is a top SNP or a fine-mapped SNP, the causal probability from FINEMAP, effect allele, effect allele frequency, sample size, corresponding gene, variant consequence, associated eQTL gene, associated eQTL associated eQTL gene and p-value in GTEx (from https://gtexportal.org/home/) [67]. See Supplementary Table S1 for detailed information about all 25 variants and Supplementary Table S2 for fine-mapping results.

Strong associations were observed between 17 SNPs in the *UNC5D* region on chromosome 8 and the occurrence of very rare functional variants in the EBV **BALF5** gene (Figures 2a, 3a), which is involved in viral DNA replication during the late phase of lytic infection. *UNC5D* is a poorly characterized gene expressed mainly in neuronal tissues, which encodes a protein that has been shown to regulate p53-dependent apoptosis in neuroblastoma cells [69]. The top associated SNP, rs2950922 (OR = 1.31, 95%-CI = 1.21-1.41, P = 4.2×10^−12^, effect allele G), is an eQTL for *UNC5D* in esophageal tissue (GTEx, [67]).

**Figure 3:**
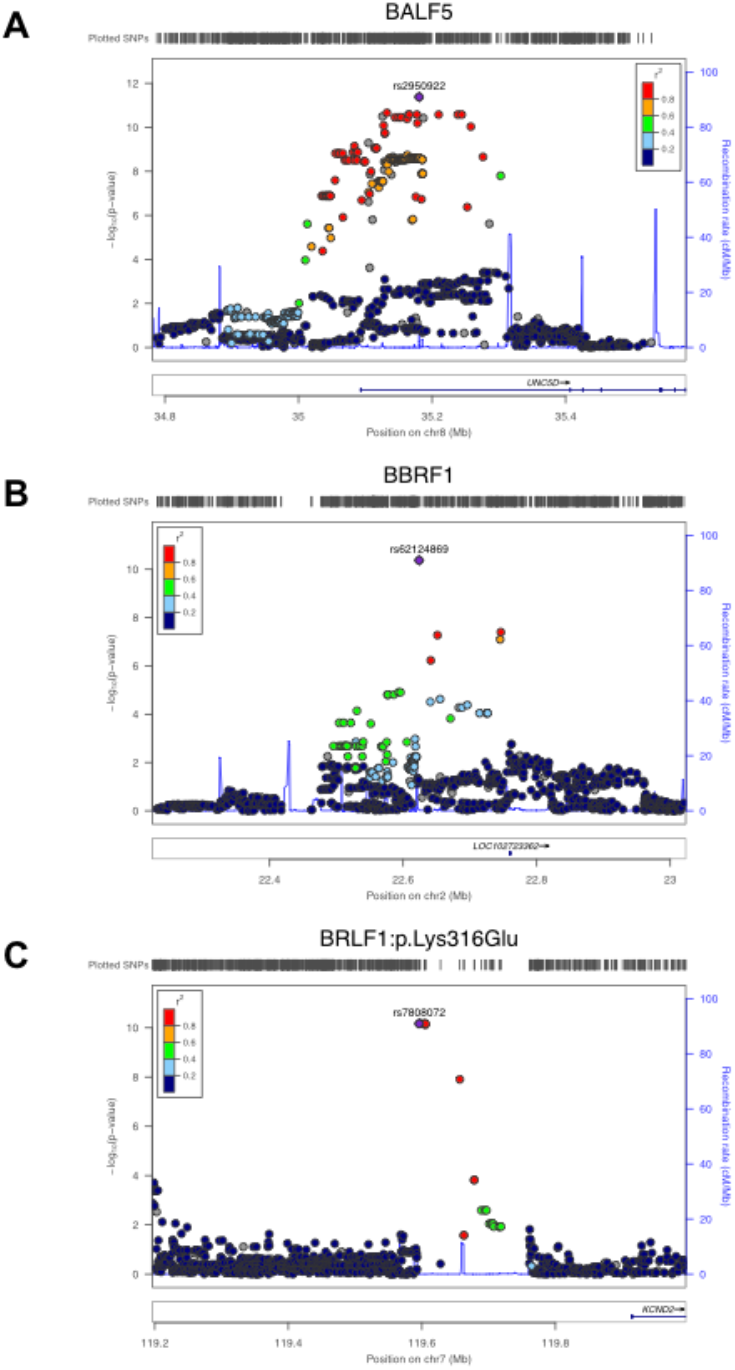
Locuszoom plots. Locuszoom plots for the three EBV association signals highlighted in red in Figure 2 (A: *BALF5*, B: *BBRF1*, C: *BRLF1*:p.Lys316Glu).

Rare amino acid variation in **BBRF1** was found to be associated with a single SNP, rs62124869 (OR = 1.29, 95%-CI = 1.19-1.39, P = 4.2×10^−11^, effect allele C), which maps to the non-coding RNA gene *LINC01830* (Long Intergenic Non-Protein Coding RNA 1830) on chromosome 2 (Figures 2b, 3b).

Finally, 7 SNPs mapping to a non-coding region of chromosome 7 were found to be associated with the EBV amino acid variant **BRLF1:p**.**Lys316Glu** (Figure 2c, 3c). The top SNP, rs6466720, had a p-value of 6.85×10^−11^ and an OR of 1.41 (95%-CI = 1.28-1.58, effect allele G). **BRLF1** controls lytic reactivation of EBV from latency and regulates viral transcription. **BRLF1:p**.**Lys316Glu** has not been described previously, but variation at the nearby residue 377 (**BRLF1:p**.**Glu377Ala**) has been shown to be prevalent in cases of nasopharyngeal and gastric carcinomas in Chinese samples [70]. **BRLF1:p**.**Lys316Glu** and **BRLF1:p**.**Glu377Ala** are in moderate LD (r2=0.55) in our dataset.

## Discussion

Because immunosuppression - and in particular T cell deficiency - favors EBV reactivation from its latent B cell reservoir, EBV viremia is frequently detected in (untreated) HIV-infected individuals with advanced disease and low CD4+ T cell counts. We hypothesized that the reactivated virus circulating in these patients could carry sequence variants acquired during primary EBV infection, thereby providing a snapshot of early adaptation to the pressure exerted on EBV by the individual immune response.

To search for a potential imprint of host genomic variation on the viral sequence, we jointly analyzed genomic information obtained from paired EBV and human samples. Viral sequence variation can be seen as an intermediate phenotype, closer to potentially causal host polymorphisms than clinically observable outcomes like viral load or disease phenotypes. As such, it allows the detection of more subtle associations, less likely to be obscured by environmental influences. In our G2G analysis, we used variation at EBV amino acid residues as outcome in multiple parallel GWAS, which allowed us to obtain effect estimations between each human genetic variant and EBV variation.

We identified two EBV genes and one EBV amino acid as associated with three regions of the human genome, spanning altogether 25 SNPs. For the GWAS with **BALF5** as the outcome, the associated human genomic region contains eQTLs for the nearby gene *UNC5D*. This gene is poorly characterized but has been shown to play a role in the regulation of apoptosis [69]. The other two EBV genes (**BRLF1, BBRF1**) provided little indication of what underlying mechanism might be at play.

Our study is limited by its small sample size and by the complexity of correcting for human and EBV population stratification. Indeed, if not carefully controlled for, the existence of population structure in the host and pathogen genome might create spurious associations or decrease real signals in G2G analyses, resulting in both type I and type II errors. With a mixed model approach and the inclusion of pathogen principal components as covariates, the genomic inflation factors of our GWAS ranged between 0.92 and 1.12. This wide range of genomic inflation factors is likely due to a combination of small sample size and complex statistical model. To prevent false positives, we adjusted for genomic inflation when extracting significant SNPs and used a conservative G2G significance threshold of 5×10^−8^ divided by the effective number of GWAS performed. Although viral genetic variation is a more precise phenotype to study than traditional outcomes, it comes at the price of decreased power due to the high-dimensional outcome. The significance threshold is thus much lower than in a single GWAS. To limit the number of statistical tests performed, we restricted our analysis to common gene and amino acid variation.

Our analyses have been performed using historical samples collected from untreated HIV-infected individuals. Considering the natural history of EBV infection in humans and its high likelihood to be acquired during the first 2 decades of life, we postulate that intra-host adaptation of EBV happened before HIV infection, i.e. with a normally functioning immune system. At the time of sample collection, all study participants had advanced immunosuppression with low CD4+ T cell counts (< 200 cells/mm^3^ of blood). We therefore assume an absence of selective pressure on EBV at that time. These assumptions limit obviously the generalizability of our findings to non-HIV-infected population. Similar studies performed during primary EBV infection or in other specific population (e.g. bone-marrow transplant recipients) would help better delineate the global impact of intra-host selection on EBV sequence variation.

Our study provides a preliminary list of statistical associations between the EBV and the human genomes. The cataloguing of the sites of host-pathogen genomic conflict is potentially useful for further functional exploration, as has been demonstrated for HIV and HCV infections. Our results require replication and validation in different cohorts and settings. Importantly, larger sample sizes will be needed to increase power and provide more robust estimations.

## Data availability

The datasets generated during and/or analysed during the current study are available in the following Zenodo repositories: G2G results are in https://doi.org/10.5281/zenodo.4289138, pathogen data in https://doi.org/10.5281/zenodo.4011995.

## Data Availability

https://doi.org/10.5281/zenodo.4289138

## Consortium

### Members of the Swiss HIV Cohort Study

Karoline Aebi-Popp^9^, Alexia Anagnostopoulos^10^, Manuel Battegay^11^, Enos Bernasconi^12^, Jürg Böni^13^, Dominique Braun^10^, Heiner Bucher^14^, Alexandra Calmy^15^, Matthias Cavassini^16^, Angela Ciuffi^17^, Guenter Dollenmaier^18^, Matthias Egger^19^, Luigia Elzi^11^, Jan Fehr^10^, Jacques Fellay^1^, Hansjakob Furrer^9^, Christoph Fux^20^, Huldrych Günthard^10^, David Haerry^21^, Barbara Hasse^10^, Hans Hirsch^22,23^, Matthias Hoffmann^24^, Irene Hösli^25^, Michael Huber^13^, Christian Kahlert^24,26^, Laurent Kaiser^27^, Olivia Keiser^28^, Thomas Klimkait^22^, Lisa Kottanattu^29^, Roger Kouyos^10^, Helen Kovari^10^, Bruno Ledergerber^10^, Gladys Martinetti^30^, Begoña Martinez de Tejada^31^, Catia Marzolini^11^, Karin Metzner^10^, Nicolas Müller^10^, Dunja Nicca^24^, Paolo Paioni^32^, Giuseppe Pantaleo^33^, Matthieu Perreau^33^, Andri Rauch^9^, Christoph Rudin^34^, Alexandra Scherrer^10,35^, Patrick Schmid^24^, Roberto Speck^10^, Marcel Stöckle11, Philip Tarr^36^, Alexandra Trkola^13^, Pietro Vernazza^24^, Noémie Wagner^37^, Gilles Wandeler^9^, Rainer Weber^10^, Sabine Yerly^38^

^9^ Department of Infectious Diseases, Bern University Hospital, University of Bern, Switzerland

^10^ Division of Infectious Diseases and Hospital Epidemiology, University Hospital Zurich, University of Zurich, Switzerland

^11^ Division of Infectious Diseases and Hospital Epidemiology, University Hospital Basel, University of Basel, Switzerland

^12^ Division of Infectious Diseases, Regional Hospital Lugano, Switzerland

^13^ Institute of Medical Virology, University of Zürich, Switzerland

^14^ Basel Institute for Clinical Epidemiology and Biostatistics, University Hospital Basel, University of Basel, Switzerland

^15^ Division of Infectious Diseases, University Hospital Geneva, University of Geneva, Switzerland

^16^ Division of Infectious Diseases, University Hospital Lausanne, University of Lausanne, Switzerland

^17^ Institute of Microbiology, University Hospital Lausanne, University of Lausanne, Switzerland

^18^ Centre for Laboratory Medicine, Canton St.Gallen, Switzerland

^19^ Institute of Social and Preventive Medicine, University of Bern, Switzerland

^20^ Clinic for Infectious Diseases and Hospital Hygiene, Kantonsspital Aarau, Switzerland

^21^ Deputy of the patient organization “Positive Council”, Switzerland

^22^ Division Infection Diagnostics, Department Biomedicine - Petersplatz, University of Basel, Switzerland

^23^ Division of Infectious Diseases and Hospital Epidemiology, University Hospital Basel, Switzerland

^24^ Division of Infectious Diseases and Hospital Epidemiology, Cantonal Hospital St.Gallen, Switzerland

^25^ Clinic for Obstetrics, University Hospital Basel, University of Basel, Switzerland

^26^ Childrens Hospital of Eastern Switzerland, St.Gallen, Switzerland

^27^ Division of Infectious Diseases and Laboratory of Virology, University Hospital Geneva, University of Geneva, Switzerland

^28^ Institute of Global Health, University of Geneva, Switzerland

^29^ Pediatria, Ospedale Regionale di Bellinzona e Valli, Bellinzona, Switzerland

^30^ Cantonal Institute of Microbiology, Bellinzona, Switzerland

^31^ Department of Obstetrics and Gynecology, University Hospital Geneva, University of Geneva, Switzerland

^32^ University Children’s Hospital, University of Zurich, Switzerland

^33^ Division of Immunology and Allergy, University Hospital Lausanne, University of Lausanne, Switzerland

^34^ University Childrens Hospital, University of Basel, Switzerland

^35^ Swiss HIV Cohort Study, Data Centre

^36^ Kantonsspital Baselland, University of Basel, Switzerland

^37^ Pediatria, University Hospital Geneva, University of Geneva, Switzerland

^38^ Laboratory of Virology, University Hospital Geneva, University of Geneva, Switzerland

## Acknowledgments

This study was supported by the Leenaards Foundation (Leenaards Prize 2015 to JF and EZ). This study has also been partly financed within the framework of the Swiss HIV Cohort Study, supported by the Swiss National Science Foundation (grant #177499), by SHCS project #743 and by the SHCS research foundation. The data are gathered by the Five Swiss University Hospitals, two Cantonal Hospitals, 15 affiliated hospitals and 36 private physicians (listed in http://www.shcs.ch/180-health-care-providers).

## Author contributions

EZ and JF conceived and supervised the work.

SR and CH performed the association analyses.

DPD, SM, JB performed EBV sequencing and curated the data.

AL prepared and analysed the viral sequencing data.

PJM, DL, ON contributed to the design of the work.

SHCS collected the samples and associated data.

SR, CH, AL and JF wrote the paper.

All authors reviewed the manuscript and approved the submission.

