## Supplementary Figures for "The influence of human genetic variation on Epstein-Barr virus sequence diversity"

Sina Rüeger[1,2#], Christian Hammer[3#], Alexis Loetscher[2,4#], Paul J McLaren[5,6], Dylan Lawless[1,2], Olivier Naret[1,2], Nina Khanna[7], Enos Bernasconi[8], Alexandra Calmy[9], Matthias Cavassini[10], Huldrych F. Günthard[11,12], Christian R. Kahlert[13], Andri Rauch[14], Daniel P. Depledge[15], Sofia Morfopoulou[15], Judith Breuer[15], Evgeny Zdobnov[2,4], Jacques Fellay[1,2,16]* and the Swiss HIV Cohort Study

### Supplementary Figures

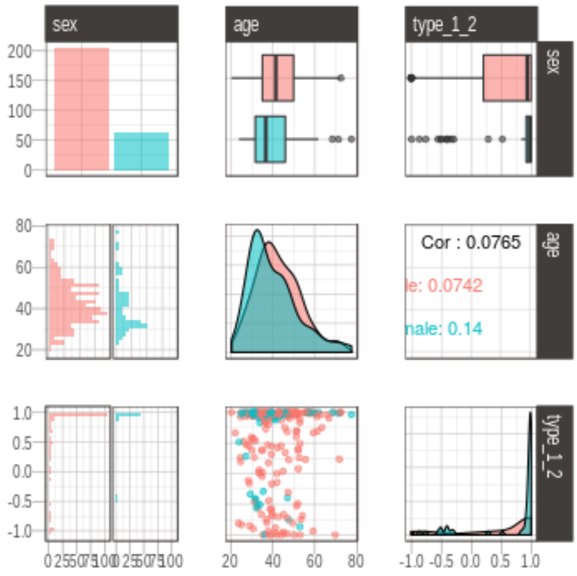


**Supplementary Figure S1:** Description of all covariates (sex, age, EBV type 1 vs 2). Each covariate is presented with all other covariates. Colour represents sex (red male, turquoise female).

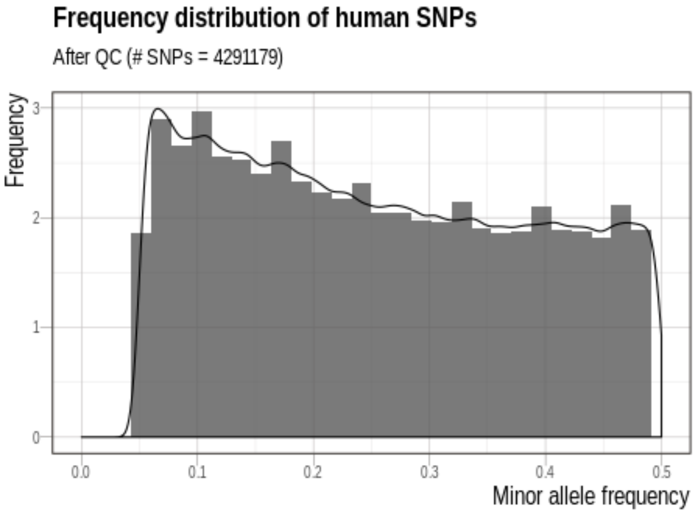


**Supplementary Figure S2:** Summary of host SNP minor frequency after QC. The x-axis represents the minor allele frequency, the y-axis the frequency.

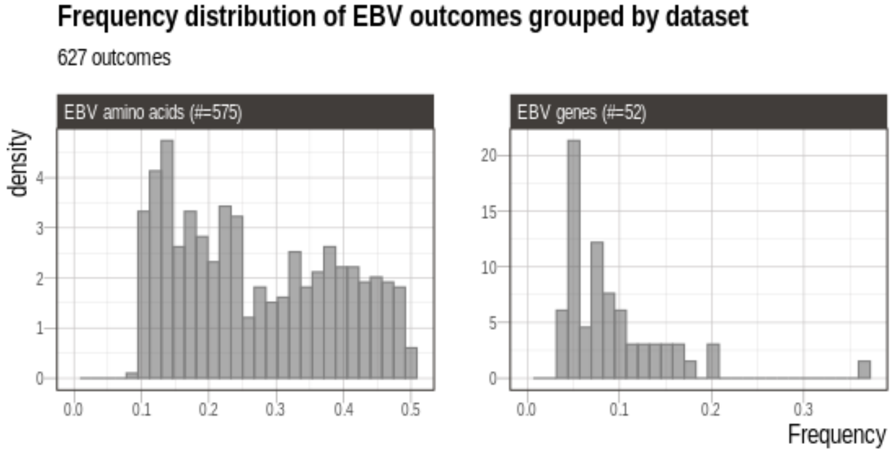


**Supplementary Figure S3:** Summary of 627 EBV gene and amino acid variants grouped into the two EBV datasets: *EBV amino acids* dataset (LHS); *EBV genes* (RHS). Each value represents the frequency for one outcome.

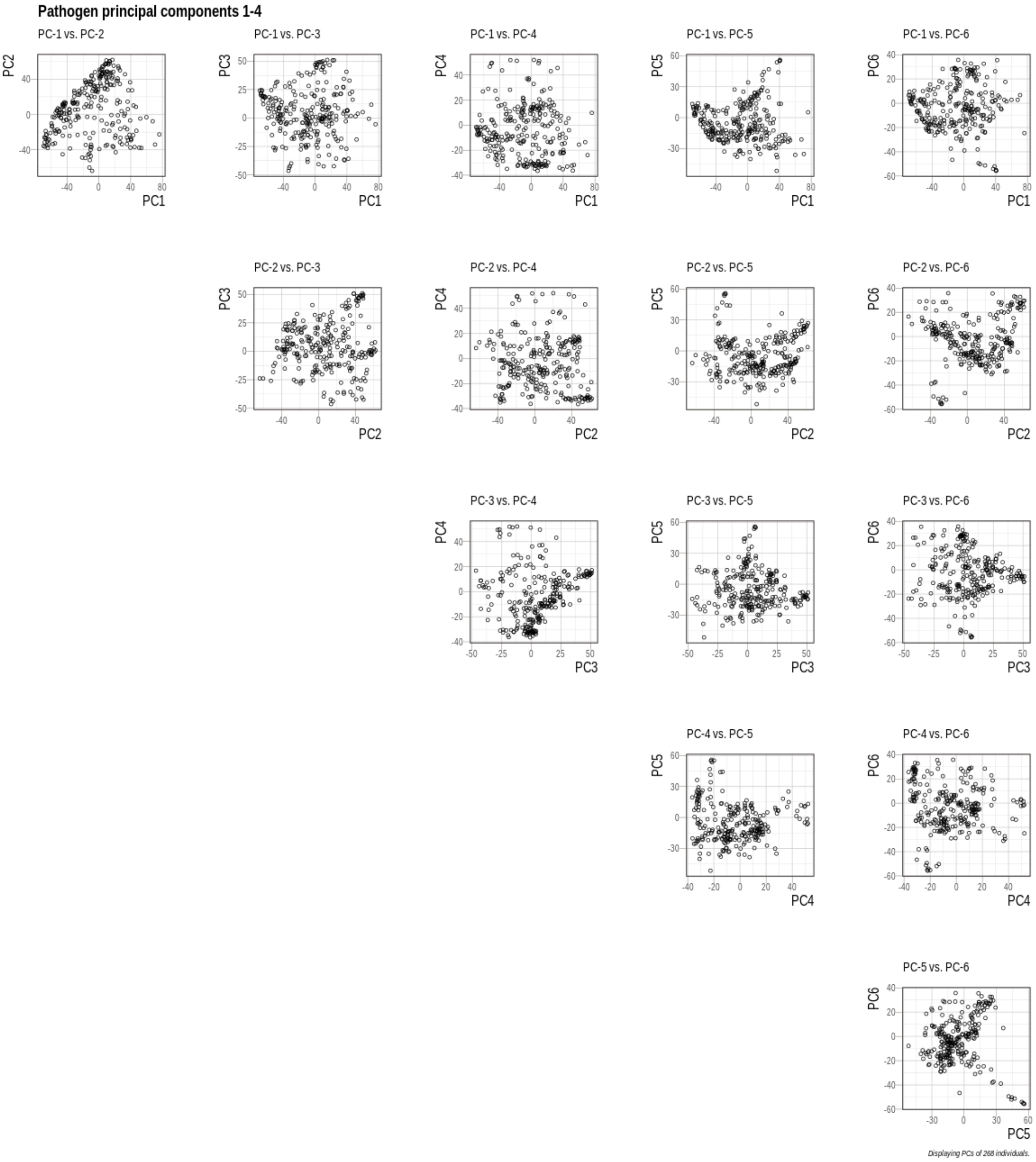


**Supplementary Figure S4:** First six pathogen principal components (PCs) plotted against each other. Each dot represents one individual. Principal components were computed based on EBV amino acid variants.

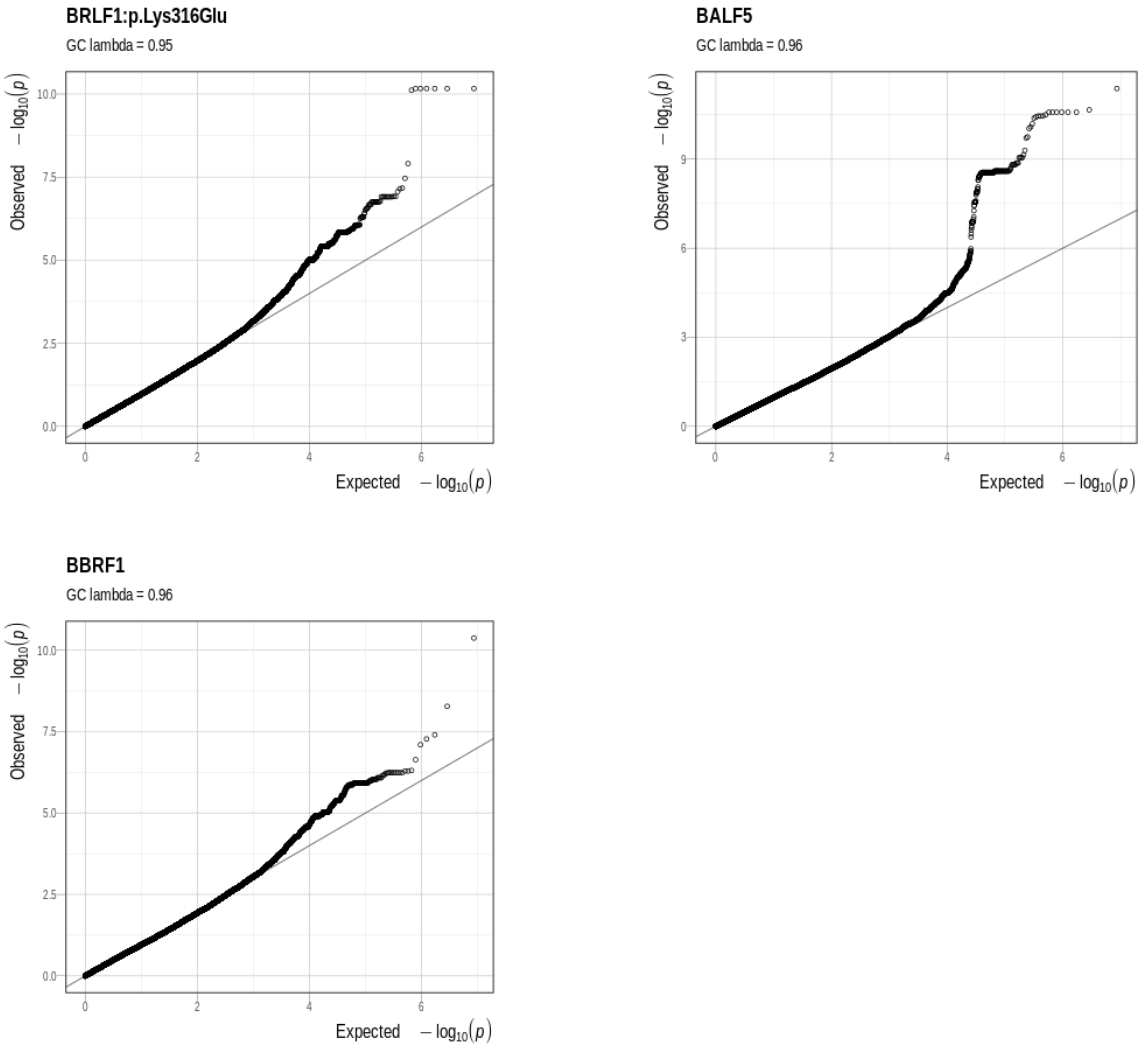


**Supplementary Figure S5**: Q-Q plots for all three EBV GWASs containing at least one G2G significant signals. Each point represents one SNP association. The x-axis displays the expected, the y-axis the observed -log10(p-value). The grey line is the identity line.

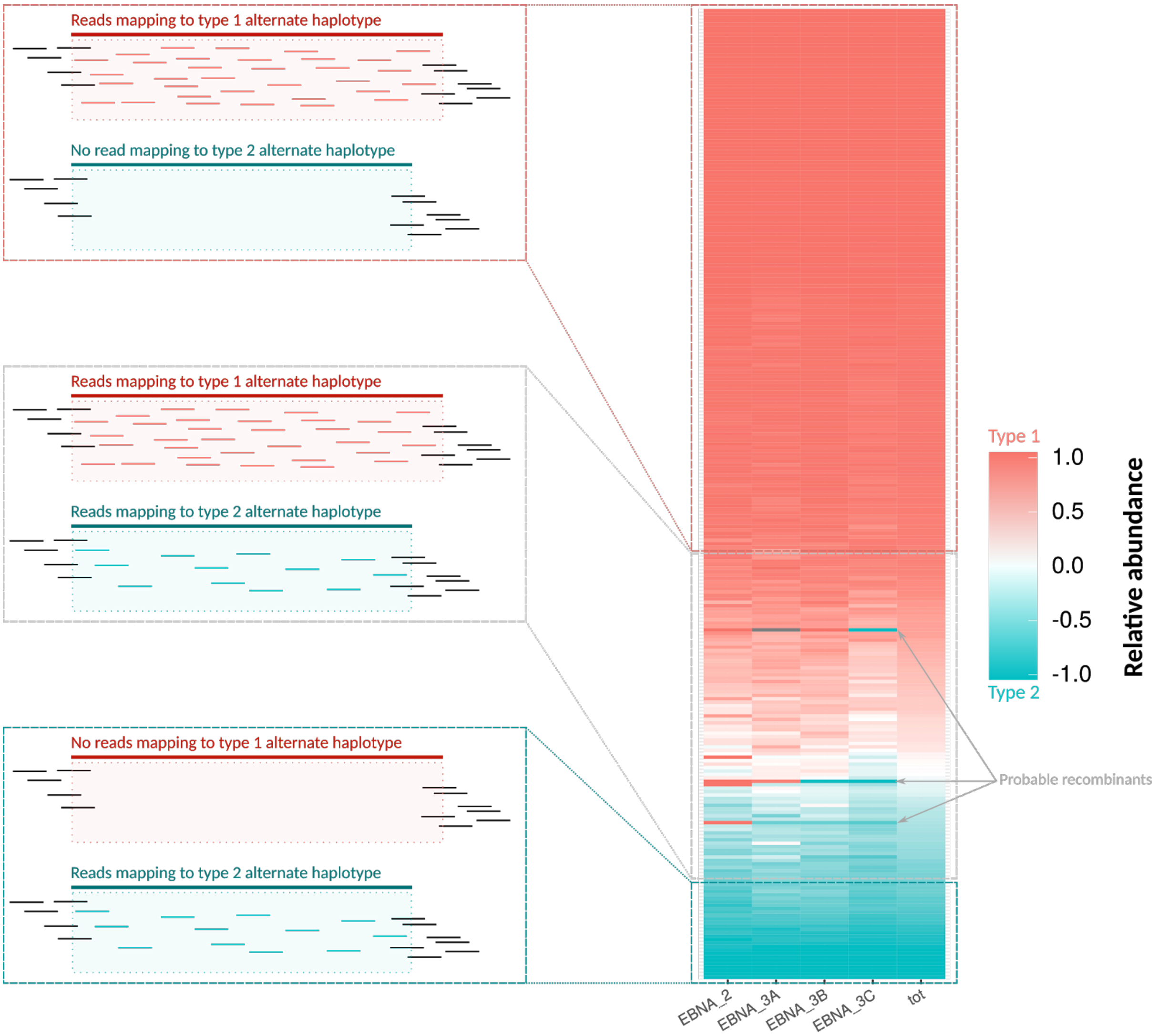


**Supplementary Figure S6**: Clonality of type 1 (red) and type 2 (turquoise) EBV strain in the blood of HIV patients. The relative abundance is calculated from reads mapping unambiguously to alternates haplotypes, as in formula 1. The heatmap shows that a substantial number of samples have mixed clonality and that 3 of them might be recombinant strains.

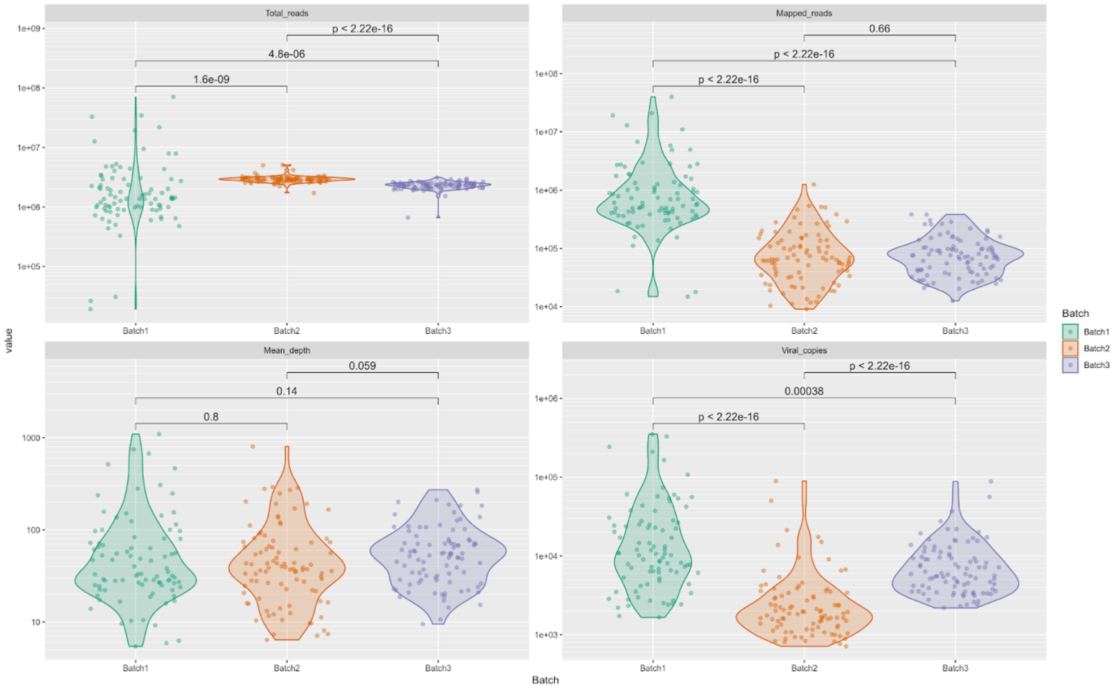


**Supplementary Figure S7a:** Statistics about coverage against AG876 and number of viral copies separated by sequencing batches. The p-values have been obtained by comparing the distributions, by batch, using the Mann-Whitney's U test.

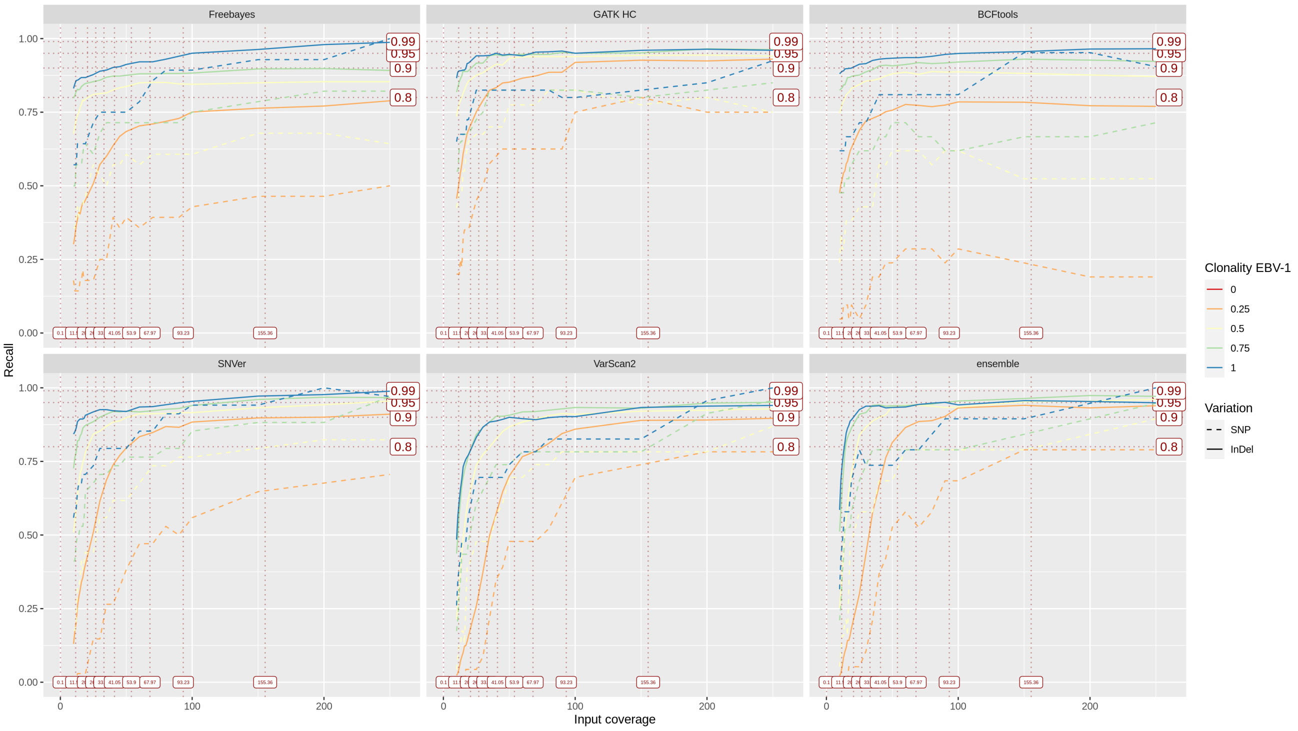


**Supplementary Figure S7b:** Performance of Freebayes, GATK HC, BCFtools, SNVer, VarScan2 against the synthetic read libraries. The ensemble plot shows the intersection of GATK HC, SNVer and VarScan2. The reads were mapped using BWA mem against AG876. The clonality (blue to red gradient) is referred as the percentage of EBV-1 reads in the libraries. The vertical axis is the recall, namely, the variant counts of a caller in a given dataset over the size of the union of all variants found in all datasets by this caller. The vertical dashed lines are the decile of the mean read depths of EBV reads in the SHCS samples.

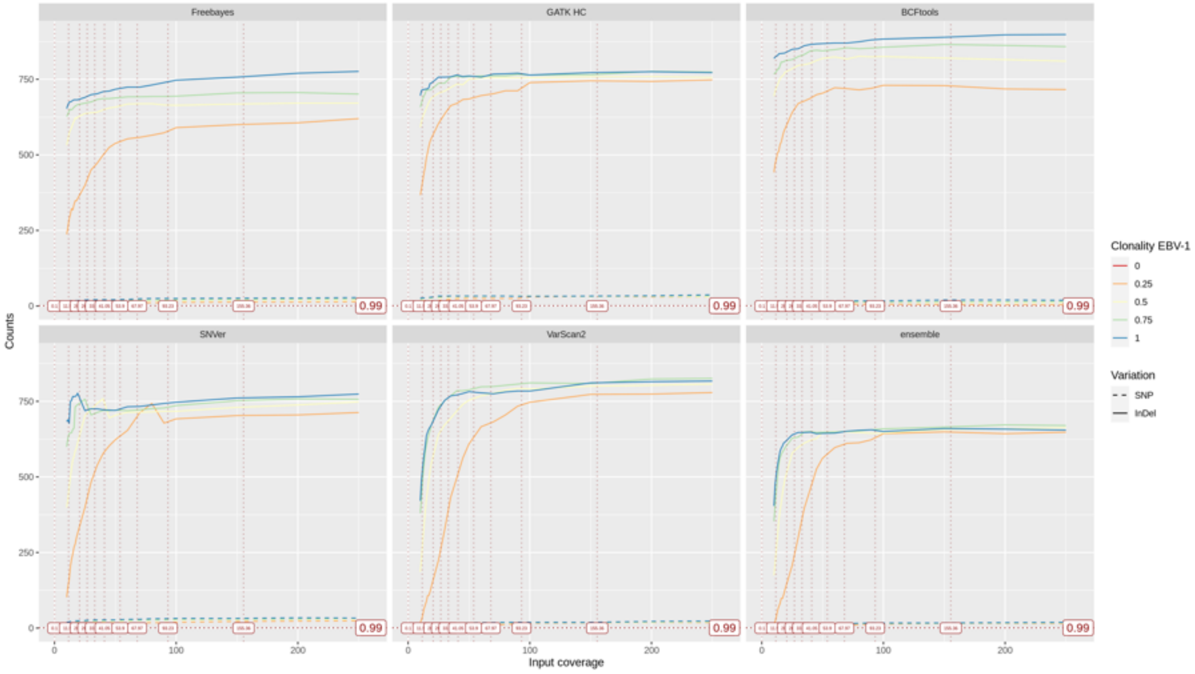


**Supplementary Figure S7c:** Raw variant counts by GATK HC, BCFtools, SNVer, VarScan2 against the synthetic read libraries. The reads were mapped using BWA mem againts AG876. The noticeable artifact observed in SNVer did not affect the result of the ensemble, let alone the result of the other callers.

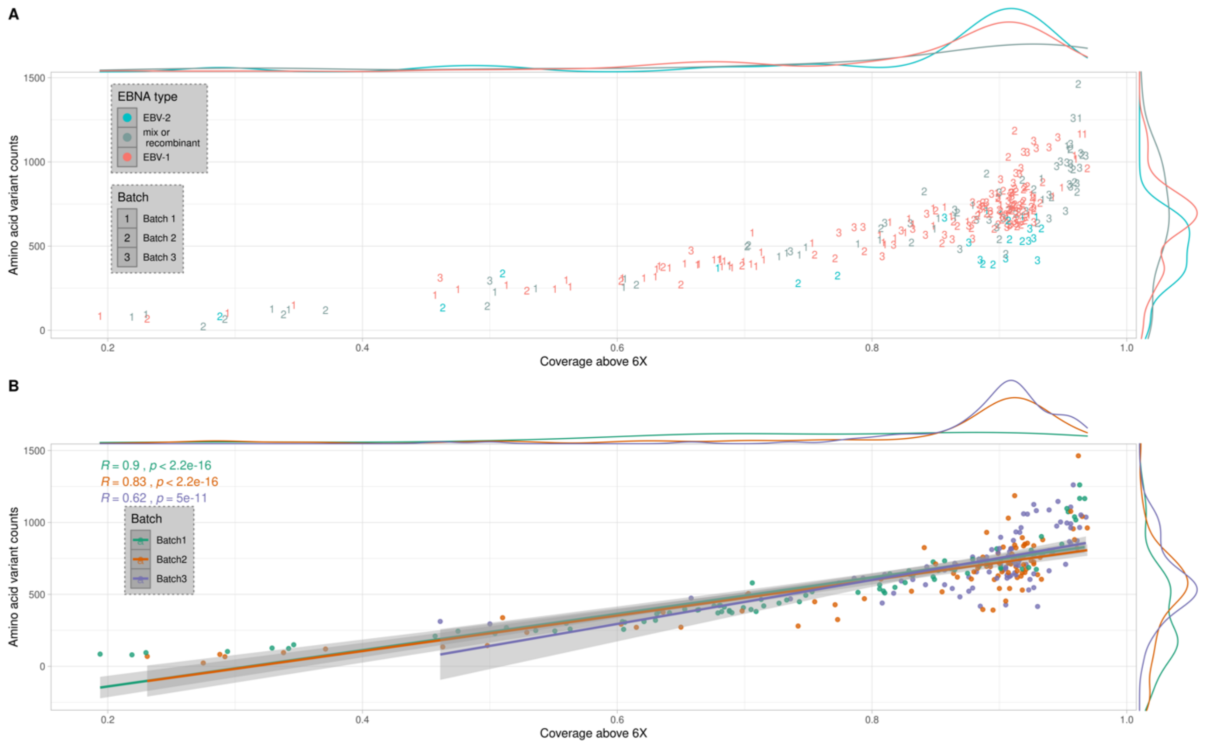


**Supplementary Figure S7d**: Correlation between coverage and amino acid variant counts. In A, the data points are colored in function of their clonality estimated by read counts against alternate haplotypes (see Supplementary Figure S6). The samples were considered mixed (gray) when the relative abundance of neither of EBV-1 (red) nor EBV-2 (turquoise) did not exceed 90%. In B, the same data is colored by batches. The results of the Pearson correlations tests between the coverage and the amino acid variant counts were colored accordingly.
